# VALIDATION OF AN INSTRUMENTED MOUTHGUARD

**DOI:** 10.1101/2022.03.02.22271563

**Authors:** Christopher M. Jones, M Rowan. Brown

## Abstract

The purpose of this study was to determine the validity of an raw and filtered acceleration time-series data from an instrumented mouthguard system against an anthropometric testing device. Testing was conducted in a laboratory using a standard impact protocol utilising the head-form of reference system and the mouthguard system. Testing occurred at 5 impact locations: facemask, front, oblique, side and back, and at four velocities (3.6, 5.5, 7.4, and 9.3 m/s) to attain a total of 55 impacts. A 160 Hz Low-pass 4^th^ order Butterworth filter was applied to the time-series data, and was reported as raw and filtered. Peak linear acceleration, peak rotational velocity and peak rotational acceleration was statistically compared to the reference measurement system. Total concordance correlation coefficient was 90.7% and 96.9%, for raw and filtered data set. Raw and filtered had intraclass correlation coefficients of 0.85 and 0.95 for peak linear acceleration, 0.99 and 0.99 for peak rotational velocity and 0.94 and 0.95 peak rotational acceleration respectively. The instrumented mouthguard displayed high accuracy when measuring head impact kinematics in a laboratory setting. The results presented in this study provide the basis on which the instrumented mouthguard can be further developed for deployment and application within sport quantify head impact collision dynamics in order to optimise performance and brain health.

## Introduction

Contact sports such as rugby union, continues to have concussion as one of the highest incidences of injury [1]. Concussion is a form of mild traumatic brain injury (mTBI) that can result from rapid acceleration or deceleration of brain tissue caused by a head impact [2,3]. While acute symptoms may persist for days to weeks following a concussion, debate continues on the potential long term sequalae as a result of repetitive head impact exposure (HIE) from concussive and sub concussive impacts, with growing concern over participation in contact and collision sport associated with long term neurodegeneration [4–6]. More recently, further evidence in an elite rugby union identified that the contact and collision events sustained over a season has been linked to an impaired redox-regulation of cerebrovascular function [7]. Therefore, the ability to quantify the cumulative HIE experienced by when participating in contact sports is imperative to develop appropriate risk metrics and understand relationships between HIE and neurocognitive outcomes [8–10].

A wide range of technology has been utilised by researchers in non-helmeted collision sports for example, head bands and skin mounted head impact telemetry systems [11,12]. However, their validity has been questioned as the sensors are not rigidly fixed to the body which can result in overestimation of recorded head impact acceleration values [8–10,13,14]. For example the work conducted by Wu et al., [15] investigated the movement of a skin mounted sensors relative to the underlying bone (soft tissue artefact) during head impact event. When compared to highspeed video it was reported that due to poor skull coupling the skin mounted sensor over-estimated peak linear acceleration (PLA) and peak rotational velocity (PRV). In comparison instrumented mouthguards (iMG) were shown to have greater validity over the other wearable technologies in accurately measuring head kinematics during an impact due to the rigid coupling of the upper dentition to the skull.

The advancement and miniaturisation of technology has given rise to the use of iMGs in the field as a means of overcoming the physical limitations associated with sensor placement and attachment [15]. There are a number of iMGs that have been used to further understand the HIE in sport [8,15–17], however there are limited validation and feasibility studies that evaluate the iMGs suitability for research and practice [2,18,19]. For example, differences observed across individual validation studies could be due to the different specification of sensors used, positioning of the sensors, and the different methodological and processing methods used by the researchers [2,8,13,17,19,20].

A recent study investigated the validation and comparison of five iMGs for measuring head kinematics and assessing brain deformation in football impacts [2] using the same methodical testing set up. The results identified that all iMG tested were valid against the industry standard anthropomorphic test dummy (ATD) for the measurement of PLA, and peak rotational acceleration (PRA). The study reported that all iMG intraclass correlation coefficients (*ICC*’s) values were above the minimum acceptability threshold of 0.80 [21,22]. Though valid further analysis with the study did report varying performances (with performance quantified as the action or process of performing a specified task or function) were reported for a number of mouthguards. For example two iMGs were unable to be utilised for machine learning models to measure brain deformation as there recorded time windows were not long enough [2,23]. In addition, one iMG reported much higher PLA mean errors of 32.4% when compared to other iMGs. A subsequent correction was published to the original paper to acknowledge that the discrepancy could be attributed to a lack of filtering of iMG kinematic data [24].

It has been well documented that the data processing of filtering can have a large effect on the measured signal and accuracy reported [6,10,19,24–27]. For example, ill-posed filters can fail in the removal of unwanted measurement artifacts but can also perturb the original signal such that discriminatory information is lost. This issue can be further exacerbated when limited to one parameter to represent the signal such as a peak value and add error to that signal through data processing techniques such as differentiation. Thus, it is essential that that the correct filter is applied to optimise and provide confidence in the filtering process. Therefore, the purpose of this study was to determine the validity of an raw and filtered acceleration time-series data from an instrumented mouthguard system against an anthropometric testing device.

## MATERIALS AND METHODS

The experimental set-up and methodology utilised is explicitly detailed in Liu et al., [2], for completeness we briefly review both below.

The experimental set-up, utilised a linear impactor and a helmeted Hybrid III ATD, both fixed securely on supporting apparatus. The iMG was moulded to fit the bespoke maxilla within the ATD head-form, to ensure tight coupling and reduce measurement errors propagated via vibration.

The ATD was equipped with a standard football helmet (Vicis Zero1), and then conducted a series of impacts to the ATD with a pneumatic linear impactor. In addition to measuring head impact kinematics with the iMG, the ATD kinematics were also measured and analysed for each impact. A set of high-accuracy sensors (linear accelerometers and angular velocity gyroscopes at the centre of gravity (CoG) of the ATD) served as the reference data (gold standard) for comparison, with the iMG obtained kinematics.

### Measurement and Specifications

The ATD head-form kinematics were measured by a triaxial accelerometer (Dytran 3273A) at the CoG as well as the three gyroscopes (DTS ARS-PRO). The accelerometer and gyroscopes measured the linear acceleration at CoG, and angular velocity respectively of the ATD. All data were acquired using the SLICE Nano & Micro software (DTS, Seal Beach, CA).

The iMG of the PROTECHT system, contained a tri-axial accelerometer (H3LIS331DL, STMicroelectronics, Genova, Switzerland) and a tri-axial gyroscope (LSM9DS1, STMicroelectronics, Genova, Switzerland). The former was sampled at 1 *kHz* (± 400 *g*, 12-bit resolution) and the latter at 1 *kHz* (±35 *rad.s*^*-1*^, 12-bit resolution). For each impact, the inertial sensors collected 104 *ms* of data for the mouthguard and 1 second for the ATD, for linear acceleration and rotational velocity. For both the ATD and iMG the trigger-point of the sensors was a linear acceleration exceeding 10 *g* in any one of the three axes. For the ATD a 300 *Hz* Low-pass 4^th^ order Butterworth filter used to remove high frequency noise. For the iMG a 160 *Hz* Low-pass 4^th^ order Butterworth filter was used to remove high frequency noise. Rotational accelerations were derived from the rotational velocity time-series using a five-point stencil approximation for the mouthguard and the central differencing method for the ATD. Peak values reported were defined as the maximum numerical value of the vector-norm of the respective time-series data. A summary of the specifications and processing is outlined below in Table 1.

**Table 1:** System specifications for the PROTECHT™ SYSTEM and ATD reference data.

|  | PROTECHT<sup>TM</sup> SYSTEM | ATD (Reference) |
| --- | --- | --- |
| Sampling rate (Accelerometer) | 1,000 Hz | 100,000 Hz |
| Sampling rate (Gyroscope) | 1,000 Hz | 100,000 Hz |
| Measurement range (Accelerometer) | $\pm 400$ g | $\pm 500$ g |
| Measurement range (Gyroscope) | $\pm 35$ rad/s | $\pm 140$ rad/s |
| Output time windows | [1, 103] ms | [-200, 800] ms |
| Output coordinate axes direction | Not parallel to standard coordinate * | J211 reference frame<br>X-front, Y-right, Z-bottom |
| Output coordinate origin | Sensor ** | Centre of Gravity (CoG) |
| Time windows after alignment processing | [-1,101] ms | [-200, 800] ms |
| Filter | Post analysis identified a 200 Hz Low pass 4<sup>th</sup> order Butterworth filter | 300 Hz Low-pass 4<sup>th</sup> order Butterworth filter |
| Derivation of Angular velocity | 5-point stencil derivative | Central difference method |

\* MG is aligned with the ATD J211 reference frame
\*\* MG inertial measurements were translated to the centre of gravity of the ATD via the rigid body transform

### Testing Protocol

Head impacts in contact sports such as football can occur at different locations and various velocities which is shown in Lui et al., [2]. Testing occurred at 5 impact locations: facemask, front, oblique, side and back, and at four velocities (3.6, 5.5, 7.4, and 9.3 m/s). Impact velocities followed the National Football League (NFL) helmet test protocol (5.5, 7.4, and 9.3 *m/s*) [28,29], with the addition of a lower velocity (3.6 *m/s*). Considering that the facemask is vulnerable to failure at repeated high-speed impacts, the facemask was subjected to only the two lower impact velocities. Additionally, due to the impact velocity being controlled by a pressurized air input, the actual velocity of the impact can be slightly different from the target velocity at times. In this study, the impact velocity error of ± 0.3 m/s was considered acceptable. The mean impact velocity and the standard deviation for all the tests were: 3.60 ± 0.13, 5.50 ± 0.08, 7.41 ± 0.08, and 9.29 ± 0.06 m/s. Finally, three repetitions of each impact location and impact velocity were conducted, which resulted in 55 impacts in total (4 locations with 4 velocities and 1 location with 2 velocities, each with 3 repetitions). For consistency, Lui et al., [2] ensured checks were performed to ensure the neck was not damaged, the chinstrap was still properly fitting, and the mouthguard had not come loose before proceeding to the next impact test [2].

### Statistical Analysis

All statistical analysis was performed using SPSS software (Version 27; SPSS Inc., Chicago, IL), with significance set at *p* ≤ 0.05. Scatterplots, and corresponding Pearson linear correlation coefficients (PLCC), coefficient of determination (R-squared), intraclass correlation coefficients (*ICC*), concordance correlation coefficient (*CCC*), a one-way analysis of variance (ANOVA) and a Bland and Altman [30] plot were used to quantify the agreement between iMGs and the reference measurement system.

Pearson linear correlation coefficients is a measure of the strength and direction of association that exists between two variables measured. R-Squared values, indicates the proportionate amount of variation in the response variable y explained by the independent variables X in the linear regression model [31]. A one-way analysis of variance is used to determine whether there are any statistically significant differences between the means of two or more independent groups identify difference between mean values of data sets. Intraclass correlation coefficients measures the reliability and validity of measurements for data that has been collected as groups [31]. Concordance correlation coefficient values were computed for the linear and rotational kinematic measures, and the combination of linear and rotational acceleration measures. The combined *CCC* value that accounts for peak linear and rotational acceleration represented the overall iMG in-laboratory validity [19]. Minimum validity threshold values for both *CCC* and *ICC* values are considered over 0.80 [19,21]. Bland and Altman plots were used in addition to R-values, *ICC* and *CCC* values as a measure of validity as Bland and Altman plots show the measurement error schematically and helps to identify the presence of heteroscedasticity [22]. This allows simple objective comparisons of validity across different measurement systems [22].

## RESULTS

Figure 1 outlines example impacts for linear acceleration, rotational velocity and rotational acceleration time series data for the ATD and the iMG (raw and filtered). Each row refers to the identical impact location but at different impact velocities .e., row (a-d), row (e-h), row (i-l), row (m-p) refer to impact velocities of 3.7, 5.6, 7.3 and 9.2 *m/s* respectively. The last column displays the Fourier frequency transformation of the raw and filtered iMG linear acceleration, rotational velocity and acceleration. Figure 2 presents Scatter plots for raw and filtered linear acceleration, rotational velocity and rotational acceleration peak values for the iMG system compared to the ATD reference system with each graph displaying the Pearson linear correlation coefficient. Statistical results are also presented in Table 2, which report the *ICC* values, *CCC* values, *R*^*2*^ values, ANOVA values, systemic bias and upper and lower limits of agreement for raw and filtered PLA, PRV and PRA for the iMG compared to the ATD reference system. Lastly, Figure 3 illustrates Bland and Altman plots comparing raw and filtered linear acceleration, rotational velocity and rotational acceleration of the iMG system against the ATD reference system.

**Table 2:** Intraclass correlation coefficients (ICC), Concordance correlation coefficients (CCC), coefficients of determination (R2), systematic bias and the upper and lower 95% limits of agreement (LOA) for PLA, PRV and PRA in the ATD, Raw iMG (rIMG) and filtered iMG (fIMG).

| | | Mean | SD | R | $R^2$ | ICC | CCC % | $p$ | Mean Relative Error % | Bias | 1.96 LOA |
| --- | --- | --- | --- | --- | --- | --- | --- | --- | --- | --- | --- |
| PLA (g) | ATD | 47.1 | 19.2 | - | - | - | - | - | - | - | - |
|  | rIMG | 55.2 | 22.3 | 0.85 | 0.85 | 0.84 | 84.8 | 0.04 | 19.7 | 8.1 | 17.0 |
|  | fIMG | 49.9 | 20.1 | 0.95 | 0.95 | 0.96 | 96.2 | 0.48 | 10.3 | 2.8 | 9.2 |
| PRV (rad/s) | ATD | 29.75 | 9.28 | - | - | - | - | - | - | - | - |
|  | rIMG | 29.74 | 9.07 | 1 | 0.99 | 0.99 | 99.7 | 0.99 | 1.9 | 0.00 | 1.48 |
|  | fIMG | 29.73 | 9.09 | 1 | 0.99 | 0.99 | 99.7 | 0.99 | 1.9 | -0.01 | 1.47 |
| PRA (rad/s<sup>2</sup>) | ATD | 2928 | 1140 | - | - | - | - | - | - | - | - |
|  | rIMG | 2852 | 1165 | 0.94 | 0.94 | 0.96 | 96.6 | 0.73 | 9.0 | -76 | 572 |
|  | fIMG | 2890 | 1153 | 0.95 | 0.95 | 0.98 | 97.6 | 0.82 | 7.6 | -38 | 481 |

Total CCC percentage for rIMG = 90.7% and fIMG = 96.9%

**Figure 1:**
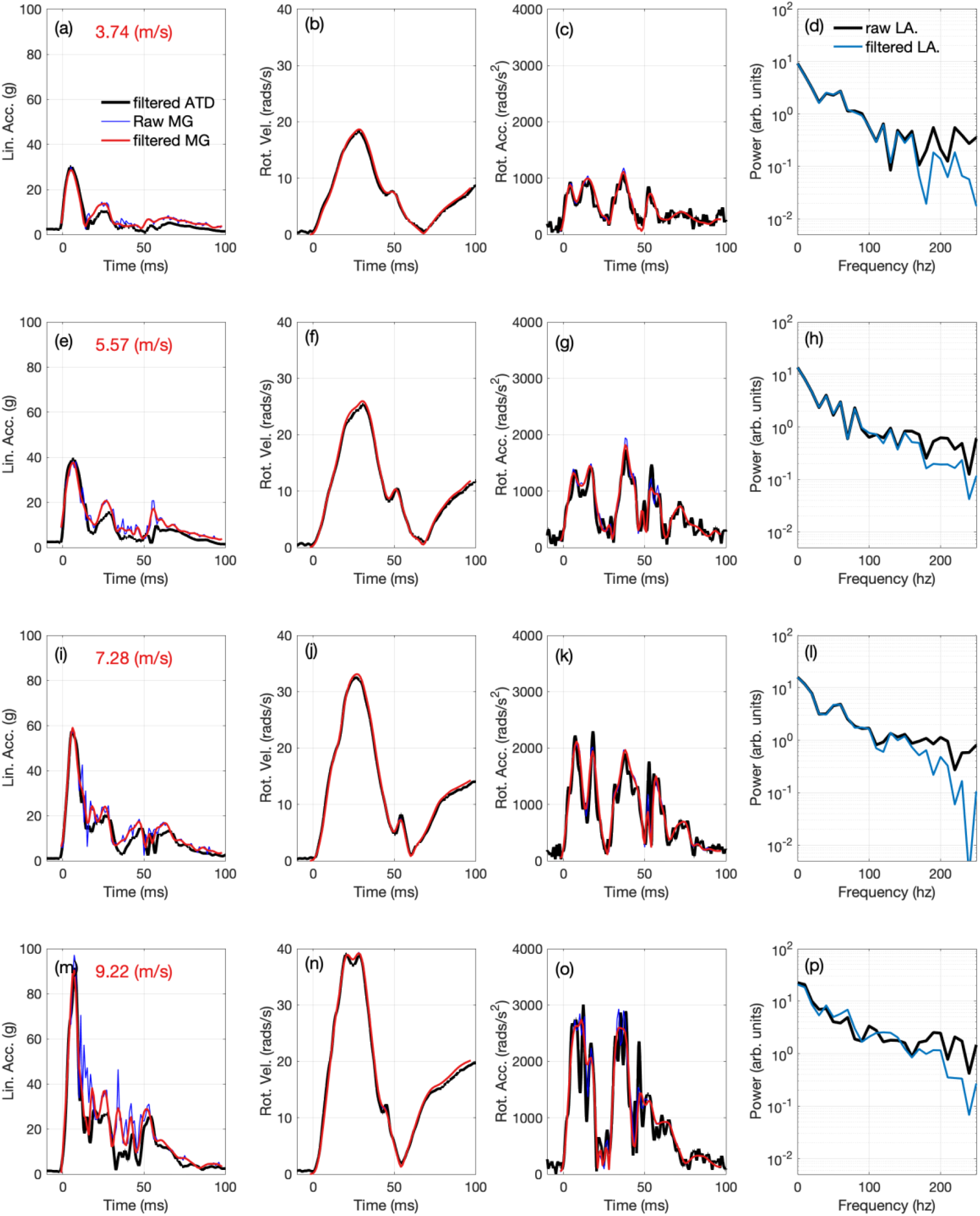
Image array of measured and numerically derived timeseries data; columns display the linear acceleration, rotational velocity, rotational acceleration (in each figure the curves corresponding to the filtered ATD and the raw and filtered iMG as shown), the last column displays the Fourier transform of the filtered iMG linear acceleration, rotational velocity and acceleration. Each row for the image array refers identical impact location but different impact velocities, i.e., row (a-d), row (e-h), row (i-l), row (m-p) refer to impact velocities of 3.7, 5.6, 7.3 and 9.2 m/s respectively.

**Figure 2:**
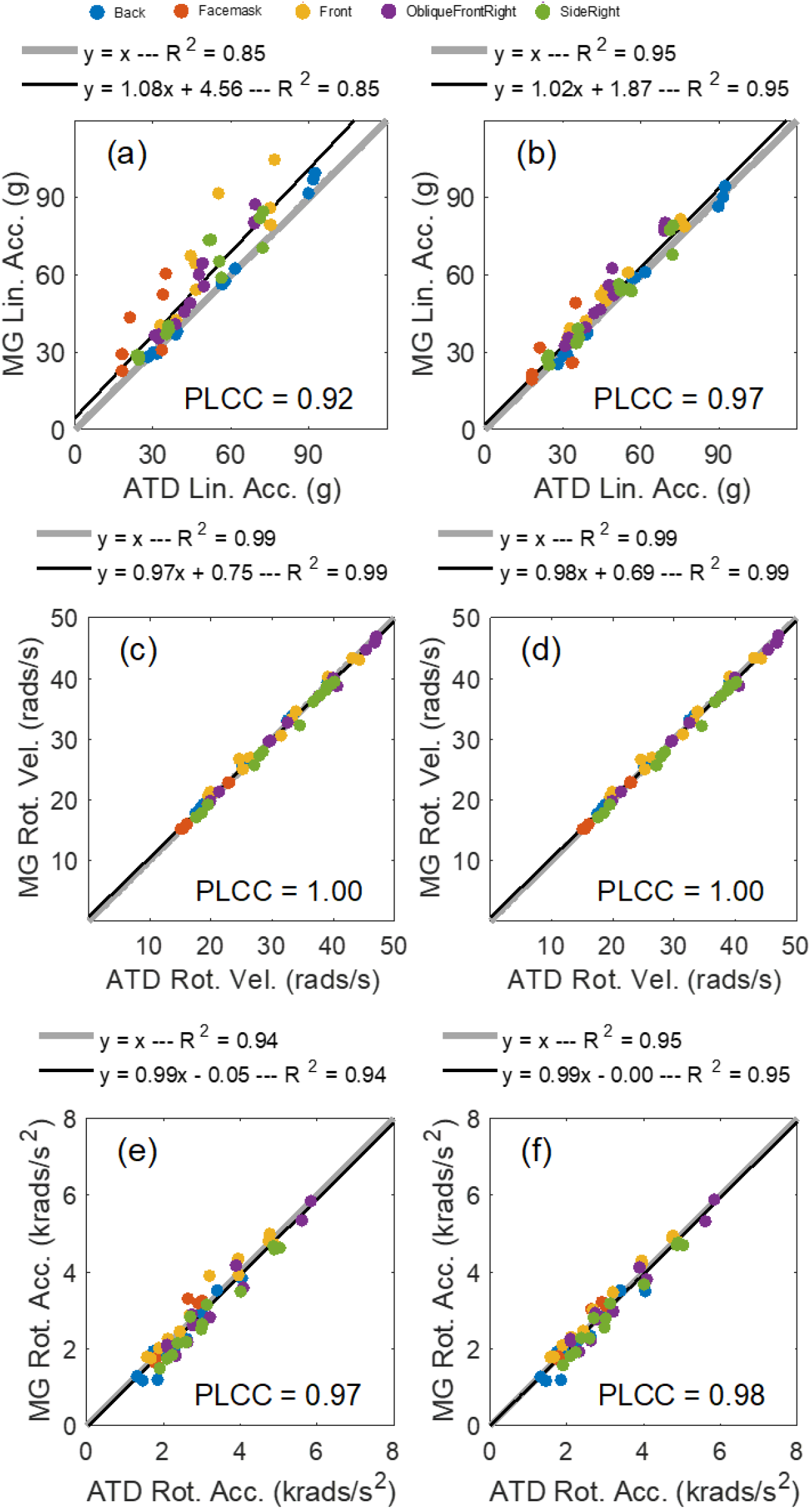
Scatter plots for raw linear acceleration (a), rotational velocity (c) and rotational acceleration (e) and filtered linear acceleration (b), rotational velocity (d) and rotational acceleration (f) for the ATD and iMG system. Each figure in the image array displays the Pearson linear correlation coefficient.

**Figure 3:**
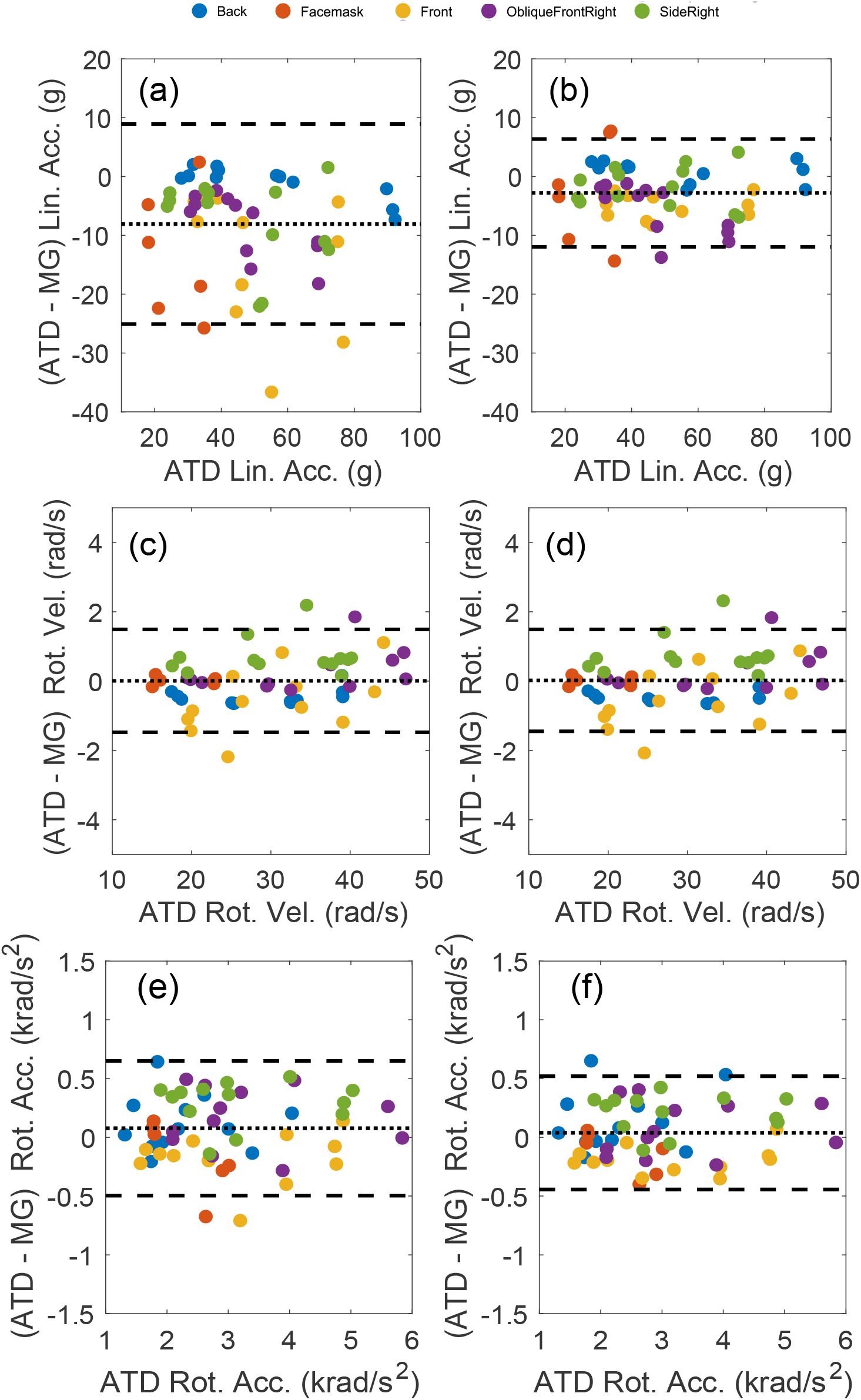
Bland and Altman (B, D, F) plots comparing raw linear acceleration (a), rotational velocity (c) and rotational acceleration (e) and filtered linear acceleration (b), rotational velocity (d) and rotational acceleration (f) of the ATD and iMG system.

## Discussion

The purpose of this study was to determine the validity of raw and filtered acceleration time-series data from an iMG system against an ATD reference system. The results showed strong positive correlations between raw and filtered iMG acceleration time-series data when compared to the ATD reference system. The measured raw and filtered iMG acceleration time-series data had total *CCC* values of 90.7% and 96.7% respectively, when compared to the ATD reference measurement system, and would have met the threshold of 80% or greater to meet a threshold for application to measure on field kinematics [19]. For total *CCC* values this study found the results of the filtered iMG of 97% to be similar, to the *CCC* values reported by Jones et al., [18] which investigated the validity and feasibility of a number of different iMG systems. The authors reported total *CCC* values of 95.3-98.3% from the highest performing iMG systems when compared with ATD reference system. Though performed within a different laboratory setting and methodical process which used a pendulum impactor on a bare headed ATD head-form in comparison to a pneumatic linear impactor on a helmeted ATD head-form. The requirements outlined by Kieffer et al., [19] in their two phase approached the action or process of performing a task or function would identity that the iMG performance in this study would be suitable to complete the second phase of evaluation [19].

No significant differences were observed for the filtered PLA, PRV and PRA values or raw PRV and PRA values with significant difference observed for raw PLA, with an 8 g difference mean which could potentially be attributed with high frequency noise observed from the methodological set up which was subsequently removed when filtered occurred. This would represent relative mean values of 19.7% decrease to 10% which is in line with values found previous studies [24].

The measured iMG and ATD data were positively correlated, with R-squared values of *ICC* values of 0.84 and 0.99 and 0.94 for raw PLA, PRV and PRA, which met the minimum acceptability for reliability and validity measures of >0.80 [21]. For example, Stitt et al., [32] found R-squared values of 0.99 and 0.99 for PLA and PRA. Furthermore, Bartsch et al., [17] reported an *R*^*2*^ of 0.99 for PLA and 0.98 for PRA and when comparing their iMG using a head impact dosimeter up to impact velocities of 8.5 *m/s*. Similarly, Camarillo et al., [33] compared a mouthguard using a custom head-form and spring-loaded impactor over eight impact velocities (2.1–8.5 *m/s*) and reported *R*^*2*^ values of 0.96 and 0.98 for PLA and PRV, respectively. Though the raw iMG values were above the threshold values for validity the application of the filter subsequently improved PLA, PRV and PRA values with reduction in relative error, improvements in total CCC values and *ICC* values of 0.96 and 0.99 and 0.95 for filtered PLA, PRV and PRA, respectively. The findings of this study are consistent with previous research that has compared the accuracy of other instrumented mouthguards for collision sports, with Greybe et al., [20] reporting *ICC* values of 0.95 and 0.99 for PLA and PRV respectively.

Bland and Altman results from this study with systemic bias values of 8.1 and 2.8 *g* for PLA, 0.00 and −0.01 *rad/s* for PRV and −76 and −38 for PRA *rad/s*^*2*^ with 95% LOA of ± 17 and 9.2 *g* for PLA, ± 1.48 and 1.47 *rad/s* and ± 572 an 481 *rad/s/s* respectively. Only two previous studies have validated iMG systems using Bland and Altman Analysis with most using only linear correlations [32]. In contrast to just causal relationship Bland and Altman Analysis enable i The results follow similar suit of those found by Greybe et al., [20] who reported a systematic bias of 2.5 *g* and − 0.5 *rad/s* and Stitt et al., [32] who reported a systematic bias of −0.49% and −1% for PLA and PRA.

One of the main findings of this stud was the improvement of results of the iMG system when filtered when compared to the ATD reference system. The main cause could be attributed to high frequency noise, which is subsequently removed through application of a 160 Hz low pass filter. This improvement is illustrated in Figure 1, with particular note identifying that higher frequency noise imparted to the iMG system as impact velocities increased. Though previous research by Greybe et al, [20] that shown no high frequency noise was apparent and subsequently did not need to be filtered, the peak linear impact magnitudes observed in that study were much lower than those observed in this study and was undcoted under different testing conditions such exhibiting impacts via a pendulum on a bare headed ATD head-from rather than a pneumatic linear impactor on a helmeted ATD head-form. It is therefore a prudent step that when research is conducted in order to validate and test iMG systems in controlled laboratory settings that the researchers conducting the experiments perform Fourier frequency analysis in order to identify and subsequently remove any high frequency noise that may occur as a result of the methodological testing set up.

With the need to accurately utilise new technology to help understand issues within sport such as concussion by measuring head impacts and more importantly using that data to understand brain deformation of the athletes in question. There is a need for a standardised testing and data handling process to ensure that the observed results are comparable and that a suitable decision can be made by researchers and practitioners about what measurement device is appropriate to use [18]. Therefore, future studies should incorporate standardised data processing techniques when validating and comparing iMG systems or using iMG systems to measure head impact accelerations.

## Data Availability

All data produced in the present work are contained in the manuscript

## Acknowledgements

The authors of this study would like to thank Dr Yuzhe Liu, Dr August Domel and Dr David Camarillo from Stanford University for data collection and permission to use this data for publication.

## Ethical approval

Not applicable

